# Disparity of secondary prevention among patients with rheumatic heart disease: A longitudinal study in Uganda

**DOI:** 10.1101/2024.03.21.24304698

**Authors:** Xinpeng Xu, Emily Chu, Hui Miao, Yuxian Du, Ryan Hoskins, Chinonso C. Opara, Neema W. Minja, Sarah Pickersgill, Jenifer Atala, Andrea Z. Beaton, Rosemary Kansiime, Chris T. Longenecker, Miriam Nakitto, Emma Ndagire, Hadija Nalubwama, Emmy Okello, Rachel Sarnacki, Jafesi Pulle, David A. Watkins, Yanfang Su

**Affiliations:** School of Public Health, Nanjing Medical University, Nanjing, China; Stanford University, Stanford, CA, United States; Harvard T.H. Chan School of Public Health, Boston, MA, USA; U.S. Data Generation & Observational Studies, Bayer Healthcare Pharmaceuticals, Whippany, NJ 07080, USA; University of British Columbia, BC, Canada; Department of Medicine, University of Washington, Seattle, WA, United States; Department of Global Health, University of Washington, Seattle, WA, United States; Kilimanjaro Clinical Research Institute, Moshi, Tanzania; Department of RHD Research, Uganda Heart Institute, Kampala, Uganda; Department of Pediatrics, School of Medicine, University of Cincinnati, Cincinnati, OH, United States; Division of Cardiology, University of Washington, Seattle, WA, United States; Division of Cardiology, Uganda Heart Institute, Kampala, Uganda; Children’s National Hospital, Division of Cardiology, Washington, District of Columbia, United States

**Keywords:** social determinants, healthcare costs, Rheumatic heart diseases, Longitudinal study, Uganda

## Abstract

**Background:** Rheumatic heart disease (RHD) is most prevalent in socially disadvantaged settings, placing a severe burden on patients and their households. The study aims to investigate the disparity of healthcare costs, including financial and time costs, among RHD patients in Uganda.

**Methods:** We enrolled 54 RHD households from the Uganda National RHD Registry between June 2019 and February 2021. The patients were interviewed in baseline and 12-month follow-up surveys. A random-effect model was applied to examine the disparity of RHD financial and time costs. Our primary outcomes are the total outpatient costs for RHD patients’ most recent visit, consisting of direct medical costs, direct non-medical costs, and time costs.

**Results:** Following the COVID-19 pandemic, the total financial cost of outpatient visits for RHD patients increased by 9 USD on average (*P*<0.01), with the change primarily driven by non-medical costs such as transportation and food (5.8 USD, *P*<0.05). Direct medical costs also increased significantly in the pandemic, with an average increase of 3.2 USD (*P*<0.1). Compared with their counterparts, non-medical costs were higher for patients with poor infrastructure, with less education, who were older, and who were male. Patients with employment experienced a higher time cost than those without (3.3 hours, *P*<0.01).

**Conclusions:** The COVID-19 pandemic significantly increased RHD outpatient costs, mainly caused by the increase in non-medical costs. Our study implies that improving infrastructure, investing in education, and providing employees with time to seek care have the potential to reduce non-medical barriers to RHD secondary prevention.

## INTRODUCTION

Rheumatic Heart Disease (RHD) is a serious but potentially preventable sequelae of Acute Rheumatic Fever (ARF), an autoimmune reaction in a small percentage of predisposed individuals to infection by Group A Streptococcus (GAS) [1]. Without treatment, ongoing valvular damage from repeated attacks of ARF results in severe disease and resultant complications, most often presenting to hospitals with heart failure in advanced stages [2]. RHD affects more than 40.5 million people globally, accounting for 1.6% of all cardiovascular deaths, an estimated 306,000 people annually [3]. Due to the widespread use of benzathine penicillin for streptococcal pharyngitis treatment and improvements in socio-economic conditions such as overcrowding and healthcare access, the global incidence of RHD declined during the latter half of the 20^th^ century and has almost been eliminated from the developed world [4–6]. However, RHD remains endemic in low- and middle-income countries (LMIC), such as Uganda, resulting in high rates of premature morbidity and mortality [7–11]. In 2018, World Health Organization (WHO) member States unanimously adopted a global resolution on RHD and ARF, for the first time officially recognizing RHD as a global health priority [12]. Furthermore, RHD is now recognized as a disease of social disadvantage [13,14].

According to the United States Centers for Disease Control and Prevention, health equity is achieved when every person has the opportunity to “attain his or her full health potential” and no one is “disadvantaged from achieving this potential because of social position or other socially determined circumstances [15].” Health inequities are often “reflected in differences in length of life, quality of life, rates of disease, disability, and death, severity of disease, and access to treatment [15].” In 2008, the WHO Commission highlighted the importance of social determinants in achieving health equity, illustrating their commitment to addressing social determinants of health [16]. RHD risk, prevention, and management are significantly impacted by social determinants of health, which creates a health equity issue in socio-economically disadvantaged settings [17]. RHD risk is correlated with social determinants such as crowding, socioeconomic status, and increased exposure to disease [17,18]. Households in LMICs often incur disproportionally high financial costs for RHD treatment, further exacerbating the condition’s heavy burden in resource-low settings. A large percentage of both direct and indirect RHD costs are paid out-of-pocket, without any reimbursement from health insurance. Direct and indirect RHD costs are also high relative to income [19,20], which is often associated with household catastrophic health expenditure [20,21]. The risk of catastrophic spending for low-income RHD patients both increases their poverty and introduces key barriers to receiving RHD treatment, resulting in an equity gap [22,23].

In Uganda, RHD poses a major health threat, with a 1-year mortality rate of 17.8% and a history of morbid complications among those with clinical RHD [24,25]. Within a representative community in the Gulu district, Uganda, data revealed that the overall adult RHD prevalence was 2.34% [26]. As in other socio-economically disadvantaged countries, overcrowding and unemployment are strongly associated with increased risk for RHD in Uganda [27]. Direct and indirect spending on RHD-related care has been shown to exert a significant economic burden in many households, driven largely by transportation, medications, and laboratory tests, whereby 20 to 35 % of Ugandan households with RHD patients experiencing catastrophic health expenditure [20].

There is robust epidemiological literature on RHD in low-resource settings, including Uganda, but a limited understanding of the disparity of RHD secondary prevention, especially at the patient level [28]. Although many studies have emphasized that social determinants impact the risk and development of RHD, there is insufficient understanding of the disparity of healthcare-related costs among patients with RHD, especially in low-income countries like Uganda. To explore the disparity of secondary prevention among patients with RHD, we conducted a longitudinal study of RHD patients and their households in Uganda and employed a random effects model to empirically analyze the disparity of RHD-related healthcare costs.

## CONCEPTUAL FRAMEWORK

Figure 1 presents how different determinants play a role in healthcare-related costs of RHD. Different determinants can impact RHD healthcare costs through several mediators. First, those with higher levels of education tend to have better hygiene practices, possibly reducing the risk of Group A streptococcus (GAS) infection and RHD progression, which is positively associated with outpatient direct costs. Patients with higher levels of education also tend to have better healthcare-seeking behavior. Secondly, given that malnutrition may lower the immune response to infection, and lower food expenditure could indicate poor nutrition intake from daily diets, we deduced that higher food expenditures could lead to a more effective immune response, thereby lowering the severity of RHD, which is related to outpatient direct costs. Third, crowding and close contact are likely to increase the spread of GAS infection, therefore increasing the risk of developing and progression of RHD, which may contribute to increasing outpatient financial costs. Finally, the accessibility to and quality of healthcare facilities varies significantly among regions within Uganda.

**Figure 1.**
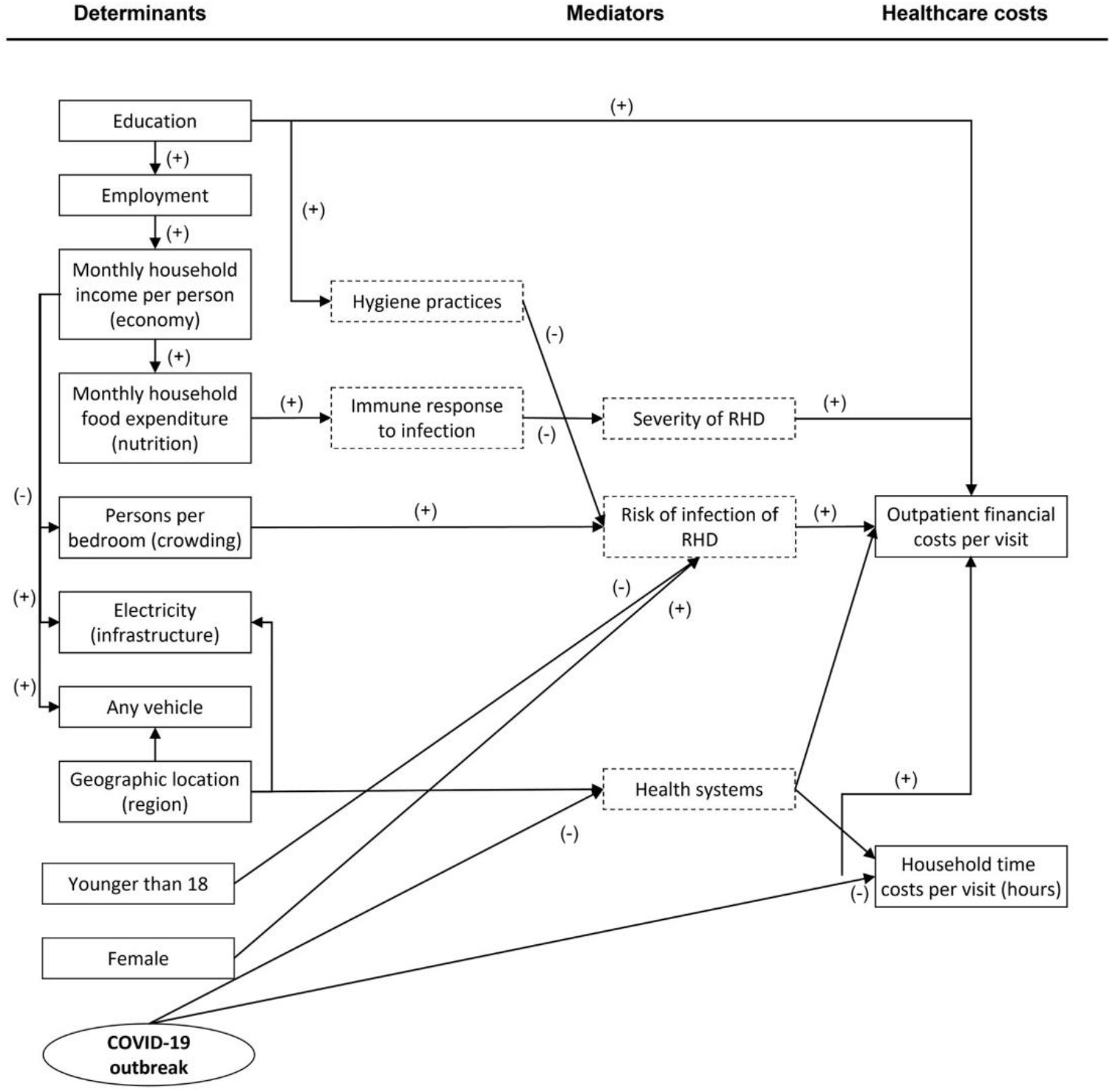
Flow-chart of determinants relating to healthcare-related costs of RHD. Note: Apart from the COVID-19 pandemic, the other social determinants in our conceptual framework are derived from a systematic review17.

The impact of the COVID-19 pandemic on health is varied. Most directly, physical limitations imposed on communities decreased patients’ access to health services at health facilities. As a result of the system-wide closure of local (primary) facilities, many patients have had to spend additional money and time going to secondary or tertiary hospitals farther from home than where they routinely received care, and at facilities where healthcare services are more expensive than those in primary healthcare sites. Disruption of the supply chain caused an increase in the costs of pharmaceuticals—costs that were passed down to patients from the health system. Additionally, social distancing measures that forced patients to work or study from home allowed for a flexible schedule that may have reduced the number of hours taken off work or school for medical appointments.

Among the determinants, some may affect other determinants in addition to the healthcare cost outcomes. A higher educational level tends to lead to better employment status and thus results in higher household income. Those with higher household income are more likely to have electricity, vehicles at home, and higher food expenditure, and are less likely to have crowding in bedrooms. Geographic location also affects infrastructure and the possibility of owning a vehicle at home. Furthermore, sex and age are regarded as risk factors for RHD. According to worldwide and regional research, the prevalence of RHD in women is consistently higher than in men [29–31]. Global burden studies reveal that the age group between 25 and 29 years old had the highest frequency of RHD in 2019 [32]. From 1990 to 2019, RHD prevalence was primarily concentrated between the ages of 15 and 49 [31]. Based on these previous study findings, we hypothesize that male RHD patients under the age of 18 may incur lower healthcare costs than female and adult RHD patients.

## METHODS

### Recruitment of Study Population

We used stratified random sampling to select RHD patients from the Uganda National RHD Registry for study recruitment [33]. Patients (age >10) who were enrolled in the RHD Registry and undergoing RHD treatment at one of three selected Regional Referral Hospitals were recruited at random. Minors participate with the authorization and informed agreement of their guardians. The three Regional Referral Hospitals (one each from the North, West, and Central regions) were selected because of the Hospital’s prior involvement in RHD-related research collaborations. There was an initial recruitment target of 1/3 of the total study participants recruited from each selected hospital.

### Data Collection

Our inquiry began in December 2018, and the investigation’s specific implementation approach is detailed in our previous study [20]. Our original study design called for the investigation of 100 patients with RHD to achieve 5% accuracy in descriptive statistics. However, due to the COVID-19 outbreak in the United States and Uganda, the Research Ethics Committee in Uganda for this study halted almost all research activities between March and July 2020. We chose to close the survey and examine the information gathered from the baseline survey due to the uncertainty around resuming research activities in Uganda. We collected data on 87 RHD patients from three regions by March 2020. In July 2020, when the Ethical Review Commission restarted research activities, we conducted a follow-up survey of patients who received a baseline survey before March 2020. The investigation duration of RHD patients varied by location. The baseline and 12-month follow-up survey times among different regions are depicted in Figure 2.

**Figure 2.**
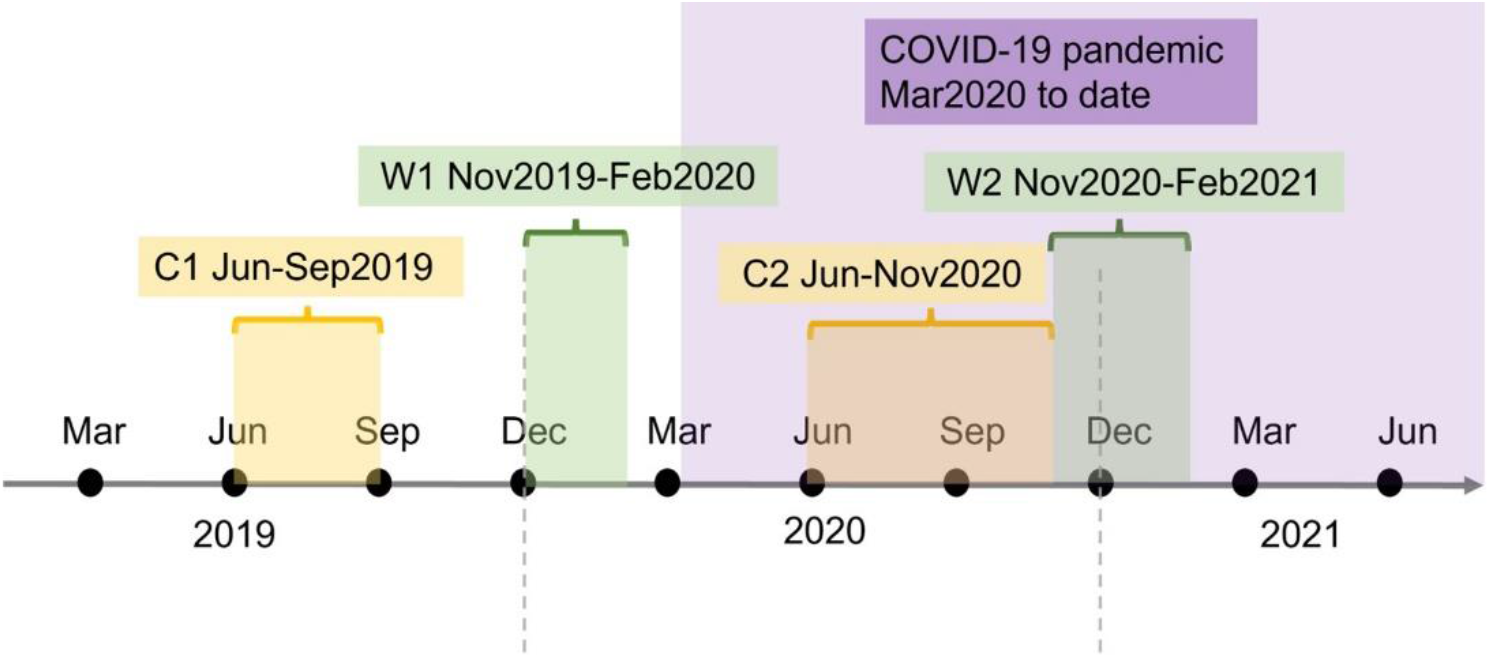
Surveys Taken in Central and Western Uganda.

Data was collected at three sites: the Northern site (Lira), the Central site (Wakiso), and the Western site (Mbarara). At the Northern site, the baseline survey was conducted from December 2018 to February 2019, followed by a 12-month follow-up from January 2020 to March 2020. At the Central site, baseline data was collected between June and September 2019, with a follow-up survey from July to November 2020. At the Western site, the baseline survey was conducted between November 2019 and February 2020, with a 12-month follow-up from November 2020 to February 2021.

We analyzed the changes in healthcare-related costs of RHD patients before and after the pandemic. Therefore, we excluded the samples originally collected from the Northern site because the follow-up data were collected before the pandemic. Our study included 108 observations from 54 RHD patients from the Western and Central regions.

### Measures

#### Healthcare related Costs

When recording patients’ healthcare costs, we recorded the expenditures generated for each outpatient record from the previous year. In the Central region, we only inquired about the patients’ latest visit during the baseline survey (Wakiso, 2019.6.6-2019.9.26), so we used the healthcare-related costs based on the currency conversion for the most recent outpatient visit as dependent variables to ensure the consistency of indicators in the study.

Direct costs associated with RHD care include both medical and non-medical costs. Medical direct costs include laboratory tests, consultations, and medicines. Non-medical direct costs comprise transportation, accommodation, and food. In addition to direct costs, we analyzed the time cost of RHD patients and their caregivers, as assessed by missed work/school hours.

#### Determinants

Pasqualina et al. (2018) summarized relevant social factors affecting the health risk and prevention of RHD in existing studies by systematic review [17]. Based on this study, we selected factors that may affect healthcare-related medical costs, including Crowding, Dwelling Facilities, Education, Nutrition, and Socioeconomic Status (SES). We used persons-per-bedroom to measure crowding, electricity as a measure for dwelling facilities, and monthly household food expenditure as a proxy for nutrition. We also included the education level of the patient, monthly household income per capita, and household vehicle ownership in the regression to control the SES of RHD patients. We also included demographic characteristics (age, sex, region) of respondents in the analysis. In addition, a dummy variable indicating before or during the COVID-19 pandemic was added to the analysis to investigate the potential effect of the COVID-19 pandemic.

### Statistical Analysis

Healthcare-related costs were first recorded in Ugandan shillings at the survey time. Then, we applied Uganda Consumer Price Index deflators to nominal baseline local currency units (LCUs) to inflate to 2020 LCUs (3.79%). We applied exchange rates based on the OANDA conversion rate of July 1^st^, 2020, to cost variables in both survey periods to produce 2020 USD (1 Ugandan Shilling equals 0.00027 United States Dollars).

We used descriptive statistics to analyze the characteristics of the RHD patients at the two survey points. Continuous variables were described by means and standard deviations. Summary statistics for categorical variables were calculated using frequency and percentage.

Finally, several panel data random effects models were used to investigate the disparity of healthcare costs among RHD patients. All statistical analyses were carried out on Stata 16.0 (Stata version 16.0, Stata Corp LLC, College Station, TX, USA).

## RESULTS

The basic characteristics of the study sample before the COVID-19 pandemic (March 2020) and after the COVID-19 pandemic outbreak started are shown in Table 1. Supplementary Table S1 provides an additional comparison of patient characteristics across all three original regions. Among the participants, a typical RHD patient is a woman over the age of 18, who has completed secondary school and is currently unemployed. After the COVID-19 pandemic began, changes in a few socioeconomic factors indicated possible hardships experienced by RHD patients, as the proportion of households without a vehicle increased from 56% to 62%, and average monthly household income per capita fell by $35.45 following the COVID-19 pandemic, more than half the average monthly household income per capita before the pandemic. Average monthly food expenditures in households fell slightly as well, decreasing by $7.21 after the COVID-19 pandemic began. The proportion of households without electricity in the sample fell from 41% to 30%, and the persons per bedroom fell from 2.57 persons per bedroom before the COVID-19 pandemic to 2.43 persons per bedroom afterward.

**Table 1.** Summary Statistics.

| Variables | Sample size | Before COVID-19<br>Pandemic Started | Sample size | During COVID-19<br>Pandemic |
| --- | --- | --- | --- | --- |
| Outpatient direct cost for the most recent visit, Mean (S.D.) | 51 | 9.15 (10.15) | 51 | 19.19 (21.04) |
| Outpatient direct medical cost for the most recent visit, Mean (S.D.) | 51 | 3.73 (7.44) | 51 | 7.06 (8.43) |
| Outpatient direct nonmedical cost for the most recent visit, Mean (S.D.) | 51 | 5.41 (4.57) | 51 | 12.13 (17.67) |
| Time cost of patient for the most recent visit (hours), Mean (S.D.) | 51 | 3.25 (4.34) | 51 | 4.07 (3.85) |
| Time cost of caregiver for the most recent visit (hours), Mean (S.D.) | 51 | 2.22 (4.54) | 51 | 2.22 (5.27) |
| Older than 18, n(%) | 54 | 43 (80%) | 54 | 46 (85%) |
| Female, n(%) | 54 | 39 (72%) | 54 | 39 (72%) |
| Primary or Less Education, n(%) | 54 | 19 (35%) | 54 | 16 (30%) |
| Unemployment Adults or Minors, n(%) | 54 | 37 (69%) | 54 | 38 (70%) |
| Without Electricity, n(%) | 54 | 22 (41%) | 54 | 16 (30%) |
| Without Any vehicle, n(%) | 54 | 30 (56%) | 53 | 33 (62%) |
| Persons per Bedroom, Mean (S.D.) | 54 | 2.57 (1.41) | 53 | 2.43 (1.17) |
| Monthly Food Expenditures (USD), Mean (S.D.) | 54 | 77.79 (58.90) | 54 | 70.58 (70.71) |
| Monthly Household Income per Capita (USD), Mean (S.D.) | 54 | 69.73 (175.07) | 54 | 34.28 (52.98) |

**Note:** Investigations were conducted on 54 RHD patients in Central and Western Uganda. There are partial missing values for some variables; therefore, the sample size is not 54.

We employed the associated costs for each RHD patient in their latest outpatient visit as the dependent variables. Supplementary Table S2 breaks down associated outpatient costs into economic and time costs for RHD patients in the three original regions. The average direct cost for the latest visit increased from 9.15 USD before the COVID-19 pandemic to 19.19 USD afterward. Average direct medical expenditures nearly doubled, increasing from 3.73 USD prior to the COVID-19 pandemic to 7.06 USD after the pandemic for the latest visit. The average direct non-medical expenditure increased significantly as well, with the direct non-medical cost more than doubling after the COVID-19 pandemic (5.41 USD vs. 12.13 USD). We also calculated RHD patients’ and caregivers’ missed work/school hours for the latest visit. After the COVID-19 pandemic, the time cost for patients of the latest visit increased (3.25 hours vs. 4.07 hours). However, the average time cost for caregivers of the latest visit remained the same (2.22 hours) both prior to and after the COVID-19 pandemic.

We used a panel data random effect model (Supplementary Table S3) to examine the association between determinants and medical and non-medical direct costs, as well as time costs.

Figure 3 presents significant associations between social demographic factors and financial costs. The COVID-19 pandemic was the significant social determinant of direct outpatient costs, direct medical costs, and direct non-medical costs, with the biggest magnitude among the 11 measured determinants. The change in outpatient costs due to COVID-19 was mainly driven by the change in non-medical costs. Five determining factors influenced non-medical costs: COVID-19, infrastructure, age, sex, and education. Specifically, households without electricity, older patients, male patients, and less educated patients experienced more transportation and food costs in seeking RHD secondary prevention. The employed had higher time costs than the counterpart. In terms of outpatient direct costs, the direct cost of the latest visit for RHD patients after COVID-19 increased by 9 USD on average (*P*<0.01). According to the breakdown analysis of outpatient direct costs, the direct medical costs for RHD patients increased by 3.2 USD after the COVID-19 pandemic (*P*<0.1). In corroboration with the trend present in outpatient direct costs and outpatient direct medical costs, the outpatient direct nonmedical cost increased by 5.8 USD following the COVID-19 pandemic (*P*<0.05).

**Figure 3.**
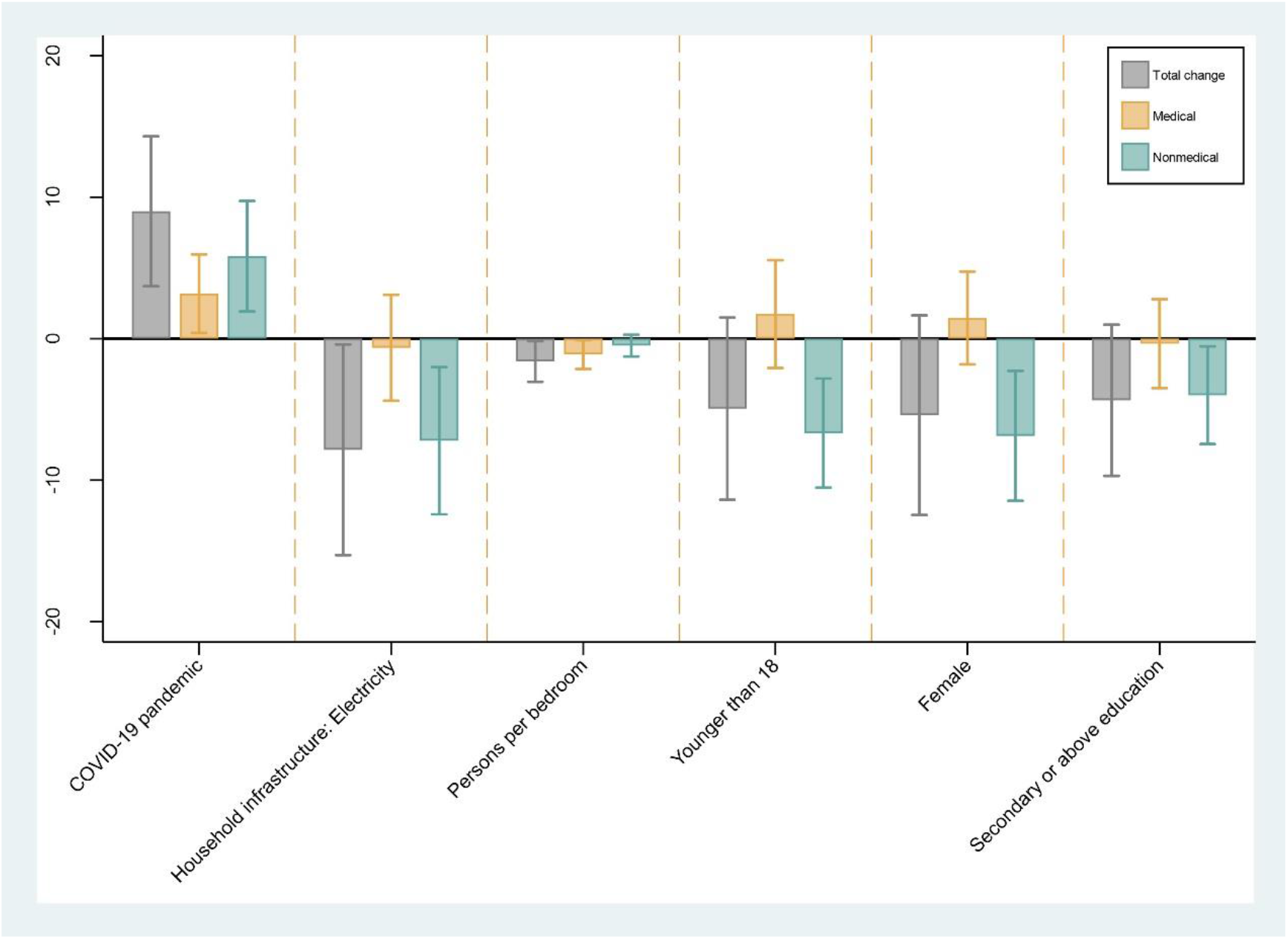
Association between social determinants and RHD financial cost: estimation from random-effects models. Note: the RHD financial cost was measured using the 2020 US Dollar.

Among other determinants, each additional person per bedroom is associated with a reduction of 1.6 USD in the direct cost for an RHD patient’s latest visit (*P*<0.1). RHD patients with household electricity spent an average of 7.9 USD less in outpatient direct costs than those without electricity in their households for the latest outpatient visit (*P*<0.1). According to the breakdown analysis of outpatient direct costs into direct medical and non-medical costs, each additional person per bedroom in the household was significantly associated with a 1.1 USD reduction in direct medical costs for the latest outpatient visit (*P*<0.1). In addition, age, sex, whether the household had electricity, and the education level of RHD patients were significantly correlated with direct non-medical costs of a patient’s latest outpatient visit. RHD patients younger than 18 spent an average of 6.7 USD (*P*<0.01) less on direct non-medical costs than patients older than 18. Female RHD patients spent 6.9 USD (*P*<0.05) less on non-medical costs than male patients. Finally, RHD patients with access to electricity spent an average of 7.2 USD less on direct non-medical costs than those without electricity in their households (*P*<0.05), and RHD patients with a secondary or higher degree in education spent 4.00 USD (*P*<0.1) less on direct non-medical costs than patients with an education level lower than secondary education. As for the time costs of RHD patients and their caregivers, RHD patients who were employed missed an average of 3.3 hours (*P*<0.01) more work or school time in the latest visit than RHD patients who were unemployed or minors. An additional 1 USD in monthly household income is associated with a reduction of 0.004 hours in the time cost for caregivers of RHD patients in the latest outpatient visit (*P*<0.1). Our study found no evidence of a significant correlation between healthcare-related costs and household monthly food expenditure, with vehicles per household, or the region dummy variable (*P*>0.1).

## DISCUSSION

We investigated the disparity of RHD secondary prevention using a longitudinal data and a random-effects panel model for RHD patients in Central and Western Uganda. Among influencing factors, we discovered that the total change in outpatient financial costs favored wealthier patients, with the lower-income population bearing a greater financial burden due to poor household infrastructure and overcrowding. External shocks, such as COVID-19, also had a dramatic effect on patients with RHD, as some faced difficult social barriers to utilizing outpatient care for RHD secondary prevention. These social barriers to treatment are mainly created by higher non-medical costs experienced by some RHD patients, particularly in transportation and food costs. Non-medical costs were higher for individuals in areas with poor infrastructure, with less education, who were older, or who were male. In addition, employed RHD patients incurred more time costs than unemployed or minor RHD patients. There is also a negative correlation between household income per capita and the time cost of the caregivers, despite the magnitude of the time cost being negligible.

As anticipated, the direct cost of the latest outpatient visits for RHD patients in our study increased significantly after the COVID-19 pandemic. Studies from other regions also consistently demonstrated that the outbreak of COVID-19 had a serious adverse impact on the healthcare of patients with non-communicable diseases. The closing of outpatient clinics in health facilities hindered patients’ access to conventional medicines and timely diagnosis. In this case, some patients discontinued treatment [34,35], increasing the risk of further deterioration of the disease [36–38], especially for patients in low- and middle-income countries [39,40]. Additionally, during the COVID-19 pandemic, social distancing policies made it more difficult for RHD patients to visit medical facilities, hence increasing their direct non-medical expenditures. The lockdowns in Uganda limited people’s access to the hospitals, with transport permits limited to emergency cases or essential workers such as health personnel and a few civil servants. Due to a lack of public transport, patient visit transportation costs increased considerably.

Patients’ direct healthcare costs have also increased as a result of the COVID-19 outbreak, which caused disruptions to the supply chain and a medicine shortage [41]. Other essential medicines have not been considered due to finances being shifted for the purchase of pandemic-related supplies. The prices of most medicines, especially those related to COVID-19 management, increased. Additionally, there has been a shift from financing other supplies to financing PPEs, which has affected medicine supplies at different facilities. Peripheral facilities are more affected compared to the regional referral hospitals. Inconvenient treatment as a result of the COVID-19 pandemic could worsen the situations of RHD patients, and a rapid fall in income could impose a heavier financial burden on RHD patients. However, the COVID-19 pandemic did not significantly increase the amount of work or school time missed by RHD patients and their caregivers due to RHD visits, likely as social distancing policies encouraged online teaching and working from home when possible.

Access to electricity is another factor associated with the reduction in the direct cost of RHD patients for the latest outpatient visit. No previous study found a significant correlation between electricity and RHD infection [17]. By using electricity as a proxy for infrastructure, our study revealed that RHD patients with electricity in their households spent 7.86 USD less than those without electricity, primarily due to a decrease in direct non-medical expenditures (7.22 USD). Patients without electricity in their households are socially and economically disadvantaged and may live in remote areas, causing them to incur higher transportation and accommodation costs during medical treatment. Where electricity is available, transportation is more convenient, and non-medical costs for RHD patients tend to lower. Electricity, and by extension infrastructure quality, are thus driving factors for a reduction in the direct cost of an RHD patient’s latest visit. However, the availability of electricity in a household has no significant effect on direct medical costs and time costs.

Crowding has been identified as a critical and necessary improvement for RHD patients [17,42,43]. From an infectious disease standpoint, living in a crowded family increases the risk of infection with GAS, ARF, and RHD [17,44]. Each bedroom in our sample housed an average of more than two persons, indicating poor living conditions and disadvantaged situations. Our study found that each additional person in a bedroom reduces the average direct medical expenditures of RHD patients by 1.6 USD, indicating the strong negative impact of overcrowding on direct medical expenditures. Persons per bedroom is a proxy variable for urbanization, and the Central region has a higher level of urbanization and population density than the Western region. Although the Western region’s economy is underdeveloped compared to the Central region, the average number of people living in a bedroom is lower. The difference between the average number of people living in a bedroom in the Western and Central regions is demonstrated by the data in the study, which reveals that the Central region had more persons per bedroom than the Western region in both the baseline and follow-up period (baseline: 2.73 vs. 2.33; follow-up: 2.58 vs. 2.20). Consequently, in terms of financial cost per patient, the average outpatient cost for RHD patients was lower in their most recent visit when there were more, rather than less, people in each bedroom, likely due to the more developed economies of urbanized regions with higher average rates of persons per bedroom. No significant rise in the time costs of patients and caregivers was observed because of overcrowding.

Studies have found that younger RHD patients and patients who are women tend to have stronger adherence to secondary prevention [45]. Our study also found that women and those under 18 years of age with RHD had higher direct medical costs than men and those older than 18 (not statistically significant), but lower direct non-medical expenditures than their counterparts, primarily driven by transportation costs. In addition, adult RHD patients who were employed faced a higher time cost associated with treatment than those who were unemployed, confirming the effect of employment on time costs associated with RHD patients. Adult RHD patients can visit the doctor and administer medication on their own, whereas juvenile RHD patients typically require a caregiver to accompany and care for them. As a result, RHD patients under the age of 18 experienced increased direct nonmedical costs and increased caregiver time costs, even though these impacts were not statistically significant. Additionally, individuals with a higher level of education were more likely to live near health facilities, providing a possible explanation for the reason RHD patients with a higher level of education have lower direct non-medical costs.

Some studies have reported a positive association between income and risk of RHD infection [46,47]. However, for RHD patients adhering to secondary prevention, our study indicated that income has no significant effect on the direct cost of the latest outpatient visit. Although a higher income is associated with a statistically significant reduction in the time cost of caregivers for the latest visit, the magnitude of the time cost is minimal and negligible. In addition, our findings demonstrated no statistically significant differences in healthcare-related costs between RHD patients living in different regions. Compared to the Western region, the Central region is more developed, has more medical institutions, and has a more concentrated medical resource distribution. The Western region is larger and has a greater proportion of rural areas. Due to limited medical resources in the Western region, RHD patients must travel to health facilities located far from their homes, which would increase their treatment and time costs. However, our study revealed that there was no statistically significant difference in costs among RHD patients in different regions, possibly due to the collinearity between region and other variables that characterize socioeconomic status, such as education, crowding, and infrastructure.

The study has several implications. To begin, the COVID-19 pandemic was the main driving factor in the increase of direct costs among RHD patients, which indicates that the current model of RHD secondary prevention delivery in Uganda is vulnerable and sensitive to pandemics. RHD patients must travel extensively to receive monthly injections, making it extremely difficult for them to receive care with lockdowns imposed. As a result, COVID-19 caused RHD patients in need of medical treatment to delay or neglect routine care. Thus, pandemic preparedness is essential for ensuring routine medical care for RHD patients in case of similar exogenous shocks in the future. In the short term, the continued decentralization of health services could mitigate the financial and time costs magnified by the pandemic. A more effective option may be to directly equip local clinic nurses with the ability to provide Benzathine penicillin G injections. In the long term, however, the development of long-acting or self-administered antibiotics must be prioritized. These methods can increase the accessibility of secondary RHD prevention for the lower-income population, and hence improve health equity among patients with RHD. Additionally, our study provides supporting evidence for enhancing infrastructure construction, improving living conditions, and promoting equality of opportunity for education in Uganda. It is also necessary to expand housing construction and maintenance, and improvements in access to amenities like clean water and electricity, as well as reductions in overcrowding, can reduce the risk of exposure to strains of streptococcus rheumatoid [48]. Finally, health education and promotion strategies are integral to improving RHD prevention and control [49]. For groups at high risk of RHD infection in particular, patient education and awareness of RHD is increasingly necessary both before and after RHD infection. Prior to infection, primary prevention can be strengthened through the dissemination of relevant health information. After infection, patient education may reduce the risk of RHD transmission, with improved education on RHD-related patient care and adherence to secondary prevention aiming to ensure the long-lasting health and safety of RHD patients.

Our study has some limitations. First, due to the impact of the COVID-19 pandemic, our investigation was halted, limiting the sample size of those included. As a result, many of our estimates are not statistically significant. Future studies are warranted for a COVID-19 pandemic analysis with a larger sample size. Second, due to the potential bias of self-reported outpatient costs this study may underestimate the actual associated costs of RHD. Finally, our study is based on the Ugandan RHD registry, which excludes RHD patients who are not included in the registry.

### Conclusions

Our study identified the disparity of secondary prevention among RHD patients in Uganda. After the outbreak of the COVID-19 pandemic, the outpatient direct costs of RHD patients increased significantly, while income decreased significantly, which brought severe economic burden to RHD patients. Importantly, household infrastructure and crowding appear to be the primary determinants of increasing direct costs for RHD patients, indicating that patients living in more impoverished conditions are burdened by higher costs. Overall, our study discovered a significant equity concern facing RHD patients in Uganda, and current data reveals a clear poverty trap: impoverished RHD patients incur greater financial burden, which lowers healthcare accessibility, further decreasing their health.

To address this poverty trap, it is essential to identify and improve these determinants, such as the COVID-19 pandemic, household infrastructure, and crowding. Primarily, to further facilitate healthcare access and reduce the financial burden of healthcare for RHD patients, measures should be taken to ensure drug access and supply in the context of the pandemic. At the same time, it is necessary to improve housing conditions and infrastructure construction for RHD patients. Finally, an emphasis on the development of self-administered or long-acting antibiotics can reduce transportation costs for RHD patients, especially during pandemic periods.

## Data Availability

The raw data supporting the conclusions of this article is available upon request to the authors, without undue reservation. Supplementary file 1: Supplementary Tables. Table S1: Household Characteristics and Demographics. Table S2: Economic and Time Costs of Outpatient Visits. Table S3: Random Effects Model Estimation Results.

## Acknowledgements

The authors would like to thank Huayi Xiong for her comments on an earlier version of this work.

## Data accessibility

The raw data supporting the conclusions of this article is available upon request to the authors, without undue reservation.

Supplementary file 1: Supplementary Tables. Table S1: Household Characteristics and Demographics. Table S2: Economic and Time Costs of Outpatient Visits. Table S3: Random Effects Model Estimation Results.

## Ethics and consent

This study was approved by the Makerere University School of Medical Research and Ethics Committee (REC RF 2018-082), the Uganda National Council for Science and Technology (SS 5081), and the University of Washington Department of Human Subjects Division (STUDY00002855). Participants provided written informed consent to participate in the study or, in the case of minors, the informed consent of their legal guardian or next of kin.

## Author’s Contributions

YS and DW conceived and designed the study. JA, RK and HN collected the data. MN and EN organized the database and supervised the data collection. XX Conducted statistical analyses. XX, EC and HM drafted the initial version of the manuscript. YD, RH, CO, NM, SP, AB, CL, EO, RS, JP, DW, and YS reviewed and interpreted the results. All authors contributed to the manuscript revision, read and approved submitted version.

